# Development and usability testing of two arts-based knowledge translation tools for parents about pediatric fever

**DOI:** 10.1101/2021.06.08.21258574

**Authors:** Shannon D. Scott, Chentel Cunningham, Anne Le, Lisa Hartling

## Abstract

Fever is defined as an elevated body temperature greater or equal to 38 degrees celsius when measured via the ear canal. It is a common bodily response in children and is typically a benign process that is self-limiting. However, fever can be an anxiety provoking event for some parents because their child can look unwell and become irritable as a result. Past attempts at translating medical knowledge about fever and its management strategies into parent-friendly formats exist; however, parent misperceptions about definition and management persist despite these educational tools.

Our research team employs patient engagement techniques to develop resources for parents to enhance the uptake of complex medical knowledge. First, our research group conducts qualitative interviews and knowledge synthesis of the literature. Following analysis, salient themes are used to develop a script and skeleton for our videos and infographics, respectively. Employing this same process, this paper discusses the development and usability testing of two digital tools for fever. Prototypes for the video and infographic were tested by parents in urban and remote emergency department (ED) waiting rooms. A total of 58 surveys were completed by parents. Overall, parents rated both the fever video and infographic favourably, suggesting that patient engaged research methods and digital formats are mediums that can facilitate knowledge transfer.

## Introduction

Childhood fever is a common symptom that presents secondary to other health conditions and is usually self-limiting [1–3]. Fever accounts for approximately 60 million clinic accesses in the United States each year and prompts numerous return emergency department visits and repeat phone calls to health care providers [1,4]. Children less than 2 years of age can experience 4 to 6 acute episodes of fever, most commonly from 3 to 36 months of age [5]. Fever plays a beneficial role in combating microorganisms and is typically benign [3–4,6]. In most cases, fever can be safely treated in the home environment with supportive therapy using antipyretics for comfort and reducing irritability rather than for fever reduction [4]. Evidence on fever literacy from the 1980s until present highlights that fever remains a stressful event for parents, as parental misconceptions still exist despite available educational resources [7–8]. Parental misconceptions about fever are outlined in a recent systematic review highlighting that parents have knowledge gaps about febrile temperature and management strategies [9].

Despite the development of numerous available resources related to fever, parents continue to have misperceptions about its definition, utility and treatment recommendations highlighting an ongoing knowledge gap [1,4,10]. Recent evidence suggests that parents continue to inaccurately define fever and associate it with harmful effects to their child (e.g., brain damage), in addition, to misunderstanding correct evidenced-based recommendations for treatment such as appropriate administration of antipyretics [1,4,9]. These challenges in the adoption, uptake and application of knowledge have been demonstrated by parents in other common childhood conditions (e.g., asthma, croup, bronchiolitis, and pain) and successfully addressed through novel patient engaged research strategies employing digital platforms in tool development such as white board animation, stories and online resources [11–13].

Translating Emergency Knowledge for Kids (TREKK) is a national program in Canada aimed at creating and disseminating evidence-based resources about acute childhood illnesses and injuries. This includes developing bottom line recommendations for healthcare providers and digital knowledge translation (KT) tools for parents and families. The Translating Evidence in Child Health to Enhance Outcomes (ECHO) and Alberta Research Centre for Health Evidence (ARCHE) research groups at the University of Alberta have collaborated with TREKK to identify and develop several KT tools for parents about common acute childhood conditions. The purpose of their research is to identify relevant evidence and develop tools that translate complex medical knowledge into easier to understand formats for parents and families. To date, ECHO and ARCHE have collaborated to develop and evaluate KT tools for a variety of childhood conditions; these are available at echokt/ca/tools.

The purpose of this report was to outline the development and usability testing of two digital KT tools (video and infographic) about pediatric fever targeted at parents. This topic was previously identified by the TREKK program needs assessment [14]. This report provides background knowledge on fever, outlines our methods of KT tool development and usability testing, presents results, and provides concluding remarks. The report will also help inform other health care professionals and researchers about our novel patient engagement research methods. Furthermore, the present research endeavour fills a unique parental knowledge gap about fever documented in the literature.

An approach to ensuring adequate parental knowledge translation involves employing patient engagement and usability evaluations on parent tools. A usability evaluation is a method adopted from computer informatics literature [15]. It involves an iterative process of gathering end-user feedback (e. g., parents) about newly produced evidenced-based tools and modifying the tools based on emerging needs before public launch [12]. The goal is to ensure that the KT tools promote enhanced knowledge adoption and are user-friendly by involving parents in their development and evaluation (i.e., through patient engagement strategies). Recently, literature has suggested that KT tools developed in a web-based format are associated with better caregiver knowledge uptake compared to traditional methods, such as pamphlets or verbal advice [16].

## Methods

A multi-phase, mixed method design employing patient engagement strategies was used to develop, refine and assess the usability of a video and infographic for parents about childhood fever. Research ethics approval was obtained from the University of Alberta Health Research Ethics Board [Pro00062904]. Additional ethics and operational approvals were obtained by individual hospitals to conduct usability testing.

### Compilation of Parents’ Narratives

Active parent and stakeholder engagement was a key focus of all stages of tool development for both the video and infographic. Narratives were informed through semi-structured qualitative interviews (**Appendix A**) and a systematic review on parental experiences with childhood fever [9, 17]. Parents of children who presented to the Stollery Children’s Hospital (Edmonton, Canada) with fever were invited to participate. Interviews (n = 15) were conducted by a research coordinator trained in qualitative methodology. Parents were asked to discuss their experiences having a child with fever. Concurrently, a systematic review was conducted to better understand previous research on parents’ experiences having childhood fever [17]. Analysis was conducted and key quotes and themes were generated and embedded into the video script and infographic skeleton. Recommendations from the TRanslating Emergency Knowledge for Kids (TREKK) Bottom Line Recommendations (BLR) for fever in young infants were also included [18]. Results from the systematic review and qualitative interviews are published elsewhere [9, 17]. Image 1 depicts the research program’s KT tool development cycle.

**Image 1.**
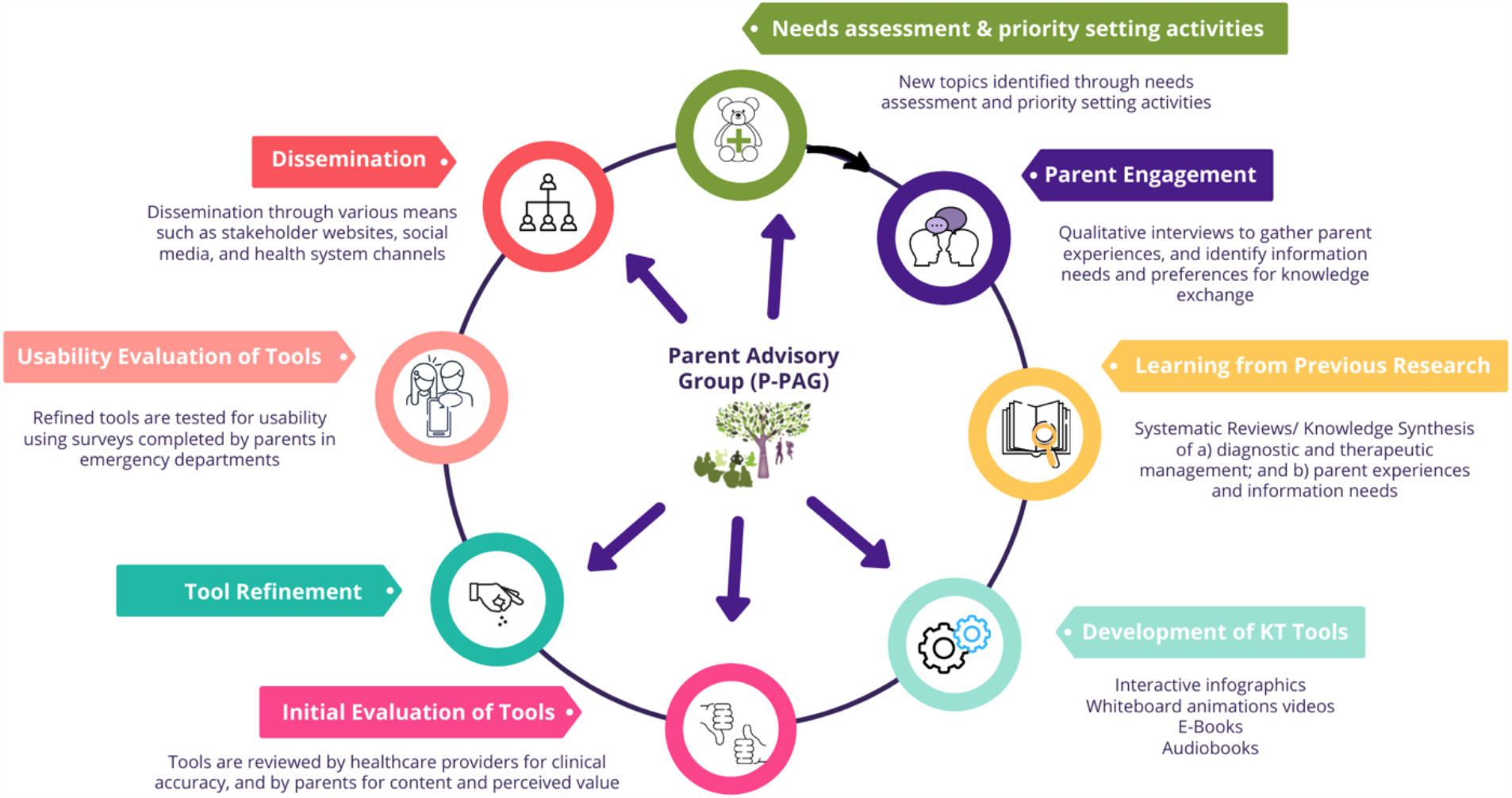
KT Tool Development Cycle.

### Prototype (Intervention) Development

Through a competitive process, we selected an animation studio with writers, digital animators, and voice actors to develop graphics, script, and prototype video. Integrating the best available evidence on pediatric fever with findings from parental interviews and systematic reviews, researchers developed a content outline (consisting of a storyboard and script), which included all information that was to be integrated into the tools. Information pertinent to management of pediatric fever was embedded within the storyline of the video, which depicted parents struggling to manage their child’s condition (**Appendix B**). Likewise, information on symptoms, treatment options, and when to seek emergency care were included in the infographic (**Appendix C**).

### Revisions

Iterative processes were used to develop the tools. Different stakeholder groups, including parents, health care professionals (HCPs), researchers, and the study team participated in several rounds of revisions. HCPs were asked to comment on the quality of information and evidence, length of the tool, aesthetics, usefulness, and perceived value. Parents from our Pediatric Parent Advisory Group (P-PAG) were asked to provide feedback on the length, stylistic elements, and highlight areas not addressed in the tools. The P-PAG meets once a month and members are asked to participate in tool development several times a year [19]. Research team meetings are held monthly to discuss the development of our tools. Feedback from these meetings is provided to developers for revisions. In comparison to other tools developed by our research programs, the fever tool was the most challenging given the lack of consensus from experts on how to best manage and treat pediatric fever.

### Video

The English-language video was 7 minutes and 4 seconds, narrated in the third person, and contained closed captioning. It portrayed two parents and their young female child’s experience dealing with fever with a health care professional’s guidance. The digital story provided reassurance that fever is a common illness. The video provides specific instructions about strategies these parents could use at home to manage fever, when to seek health care provider assessment and, details about what will happen once a child their health care provider with a fever. Screenshots from the video are included in **Appendix B**. The video included closed captioning.

### Infographic

The infographic is a web-based tool that allows the user to choose what information they wish to view and functions in the same way as a webpage, allowing parents to scroll through the page. The graphics relate to the evidence-based content embedded throughout the tool and include illustrations of children, parents and management techniques. Parents or caregivers are able to scroll and click on the information to choose what content they need. The infographic website is comprised of six sections, which include “Home (describes what a fever is)”, “Choosing a Thermometer”, “Measuring Temperature”, “When to Seek Help”, “Home Care Tips”, and “Resources”. Screenshots from the infographic are included in **Appendix C**.

### Sample and Surveys

Parents were approached for recruitment to participate in an electronic usability survey in two Canadian Emergency Department (ED) waiting rooms representing urban and remote health regions (Stollery Children’s Hospital in Edmonton, Alberta; Stanton Territorial Hospital in Yellowknife, Northwest Territories). Members of the study team approached the parents in the ED for recruitment. Upon consent, the survey software would randomize which tool (e. g., video vs infographic) the participant viewed. After parents viewed either tool, the usability survey was administered. Usability testing exists to inform if a tool meets its intended purpose by end users, in this case parents. Our usability survey was informed by a systematic review of over 180 usability evaluations and comprised of a 5-point Likert items assessing: 1) usefulness, 2) simplicity, 3) level of engagement, 4) satisfaction, 5) quality of information, 6) perceived value (**Appendix D**) [14]. Likert items ranged from strongly agree (5) to strongly disagree (1). At the end of the survey, parents were also offered to provide their positive and negative feedback of the tool via two free text boxes. Study team members were available in the ED to provide technical assistance and answer questions as parents were completing the surveys.

Data were collected using the SimpleSurveys platform on iPads. Data are stored on a secure, Canadian server. iPads were optimized with rigorous security features, including passcode login, data encryption, GPS tracking, and remote wipe capability.

### Data Analysis

Survey data were cleaned and analyzed using Microsoft Excel and SPSS v. 24 (Corp I., IBM SPSS Statistics for Windows). Descriptive statistics (e. g., frequencies) and univariate analysis (e. g., measures of central tendency) were generated for Likert responses. Independent sample t-testing was conducted to compare the usability of the video vs the infographic. Open ended survey data were analyzed using thematic analysis [20].

A timeline of the development of this tool can found in **Appendix E**.

## Results

58 parents from 2 ED waiting rooms waiting rooms participated in the usability survey. Of those, 31 reviewed the infographic and 27 reviewed the video.

A total of fifty-eight surveys were completed by parents from two sites. Twenty-seven participants viewed and completed the survey on the fever video, and 31 parents viewed and completed the infographic specific survey. Usability measures were assessed using a 5-point Likert scale with 1 being the lowest (i.e., most negative) and 5 being the highest (i.e., most positive) response. The overall comparison results of the usability survey measuring each tool are presented in table 2. Statistical analysis included descriptive statistics and means of central tendency and t testing compared Likert scale responses between tools.

**Table 1.** Demographic characteristics of parents who assessed the usability of the fever e-tools (N=58)

| Characteristic | n (%) |
| --- | --- |
| Sex |  |
| Female | 44 (75.9) |
| Male | 14 (24.1) |
| Age |  |
| 20-29 years | 7 (12.1) |
| 30-39 years | 23 (39.7) |
| 40-49 years | 17 (29.3) |
| 50-59 years | 9 (15.5) |
| 60 years and older | 2 (3.4) |
| Marital Status |  |
| Married/Partnered | 42 (72.4) |
| Single/Separated/Divorced/Widowed | 14 (24.1) |
| Missing | 2 (3.4) |
| Education |  |
| Completed primary school | 1 (1.7) |
| Some high school | 6 (10.3) |
| High school diploma | 4 (6.9) |
| Some post-secondary | 6 (10.3) |
| Post-secondary certificate/diploma | 20 (34.5) |
| Post-secondary degree | 11 (19.0) |
| Graduate degree | 8 (13.8) |
| Other | 1 (1.7) |
| Household Income |  |
| Less than \$25,000 | 6 (10.3) |
| \$25,000-\$49,000 | 6 (10.3) |
| \$50,000-\$74,000 | 9 (15.5) |
| \$75,000-\$99,000 | 9 (15.5) |
| \$100,000-\$149,000 | 9 (15.5) |
| \$150,000 and over | 12 (20.7) |
| Prefer not to answer | 6 (10.3) |
| Number of Children in the Family |  |
| 1 | 14 (24.1) |
| 2 | 17 (29.3) |
| 3 | 11 (19.0) |
| 4 | 8 (13.8) |
| 5+ | 8 (13.8) |

**Table 2.**
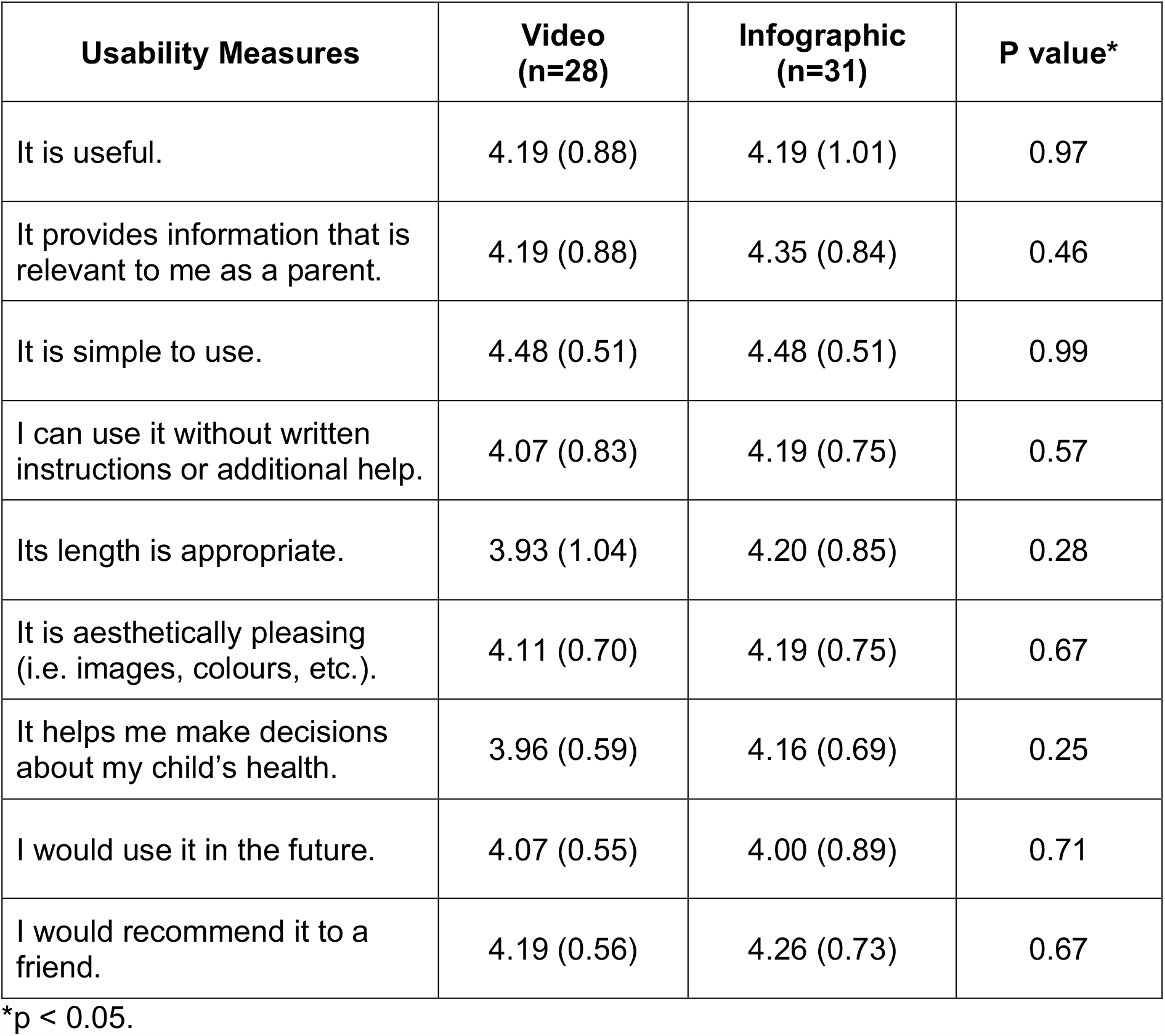
Means (SD) of participant responses to the usability survey (n=58 participants)

Overall, parents rated both tools positively with most parents answering agree and strongly agree to all items. No statistically significant differences were found between tools. When converted to numerical values, we obtained a mean of at least 4.07 out of 5.00 for both the video and the infographic. Combined frequencies reported 52 parents selecting agree (n=29) or strongly agree (n=23) when asked if it was “useful” (video mean = 4.19, SD = 0.88, infographic mean = 4.19, SD = 1.01). Parents also felt that both tools were “relevant” (video mean = 4.19, SD = 0.88; infographic mean = 4.35, SD = 0.84) as most parents responded with agree (n=28) and strongly agree (n=25). When parents were asked if they could use either tool without instruction (video mean = 4.07, SD = 0.83; infographic mean = 4.19, SD = 0.75), most parents responded with agree (n = 36) and strongly agree (n = 17). Parents scored the length of the tool slightly lower for the video as compared to the infographic (video mean = 3.93, SD = 1.04; infographic mean = 4.20, SD = 0.85), however, this difference was not statistically significant. Parents rated both tools as aesthetically pleasing (video mean = 4.11, SD = 0.70; infographic mean = 4.19, SD = 0.75). Parents responded with higher scores for future use of the infographic as compared to the video, but the difference was not statistically significant (video mean = 3.96, SD = 0.59; infographic mean = 4.16, SD = 0.69). Both the video and infographic also scored favorably in the category of “helping me make decisions about my child’s health (video mean = 4.07, SD = 0.55; infographic mean = 4.00, SD = 0.89). As well, both tools were highly recommended by parents (video mean = 4.19, SD = 0.56; infographic mean = 4.26, SD = 0.73). Table 2 summarizes the results for each usability measure for both tools.

Open ended responses were limited and as such, could not be analyzed using qualitative methods as planned. Feedback was mainly positive, with parents commenting on how easy the information provided was to understand and the graphics and visuals of both tools. When asked about negative feedback, parents indicated that there were “none” or stated, “nothing negative to say”.

## Conclusions

Overall, the usability testing demonstrated positive results for both digital tools. There were a wide variety of parents recruited from both rural and remote sites as shown in our demographics table. There were some very subtle differences in the raw scores for length and future use for the video, however, these differences were not found to be statistically significant. Open-ended responses were also found to be positive and no refinements were made to either tools based on our results. Overall, parents scored each tool favorably with “agree” or “strongly agree” to all usability measures.

Our results suggest that digital parental knowledge translation tools that are developed using patient engaged methods produce tools that are highly rated by the end users, are considered useful for parental decision-making, and contain relevant information tailored to parent knowledge needs and experiences. Participants in the usability testing found both tools to be engaging and informative.

Developing digital knowledge translation tools using an iterative process is an essential step in KT tool development aimed at pediatric families. As previous literature has outlined, these tools are essential to reducing misperceptions parents may have about fever, and to provide clear and evidence-based management strategies. Our usability findings are important as past literature has suggested that despite educational materials parents still struggle to understand fever with conventional resource development methods [3–4, 7, 21–22].

**The tools can be found here: http://www.echokt.ca/tools/fever/**

Note: Our KT tools are assessed for alignment with current, best-available evidence every two years. If recommendations have changed, appropriate modifications are made to our tools to ensure that they are up-to-date.

## Data Availability

Data are securely stored at the University of Alberta and only accessible by study team members to protect participant privacy and confidentiality.

## Other Outputs from this Project

## Appendices

### Appendix A – Interview Guide

Parents will be interviewed to understand their experience having a child with fever. Semi-structured interviews will be conducted with parents in order to get their “narrative” or experiences. The following questions will be used to guide these interviews. Being true to semi-structured interview techniques, interview questions will start broad and then move to the more specific.

1. Tell me about your experience having your child experience fever.
2. Tell me about your child that was ill. How old is your child? How was your child ill? Has your child previously had fever?
3. How did you feel during this experience?
4. What did you do to manage symptoms of fever? (any techniques you used, for example, giving Tylenol, talking with family/friends, etc.)
5. What strategies were put in place by health care professionals to help your child? (for example, giving/prescribing medication). Did they ask you to do anything?
6. How did your child manage the experience? How did you feel about the outcome of this situation?
7. If presented with the same situation again, would you do anything differently? If so, please tell me.

### Appendix B – Video

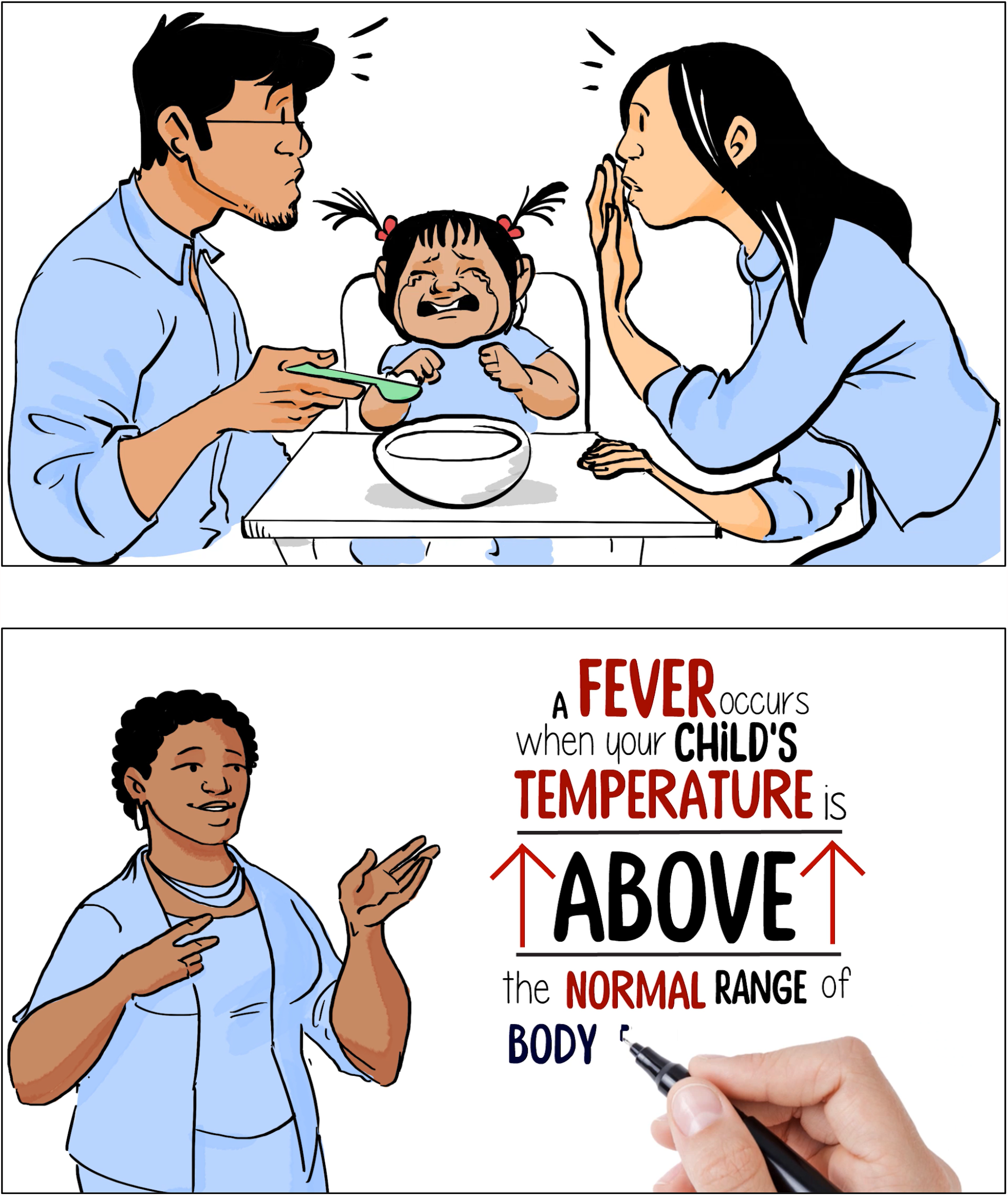

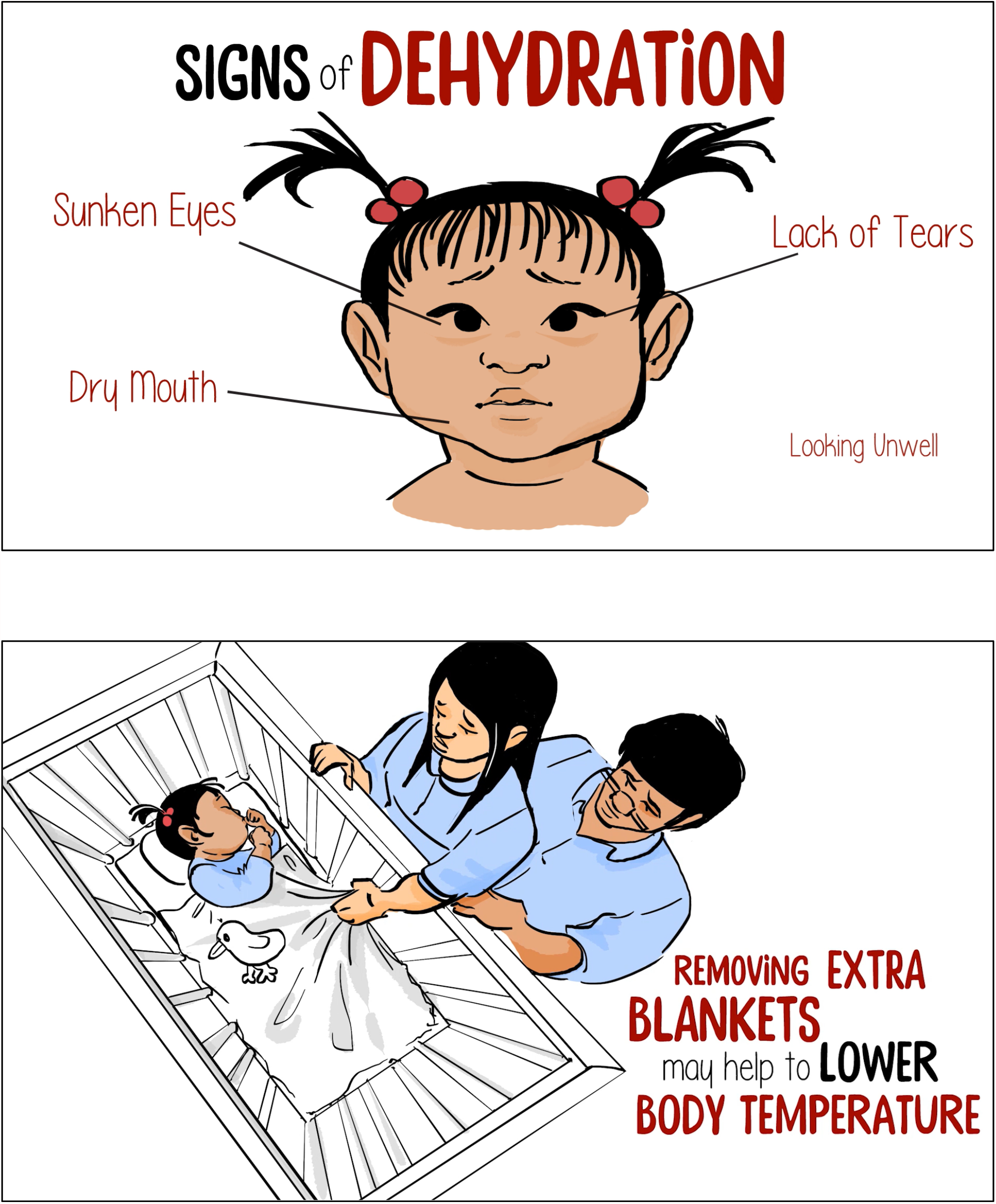

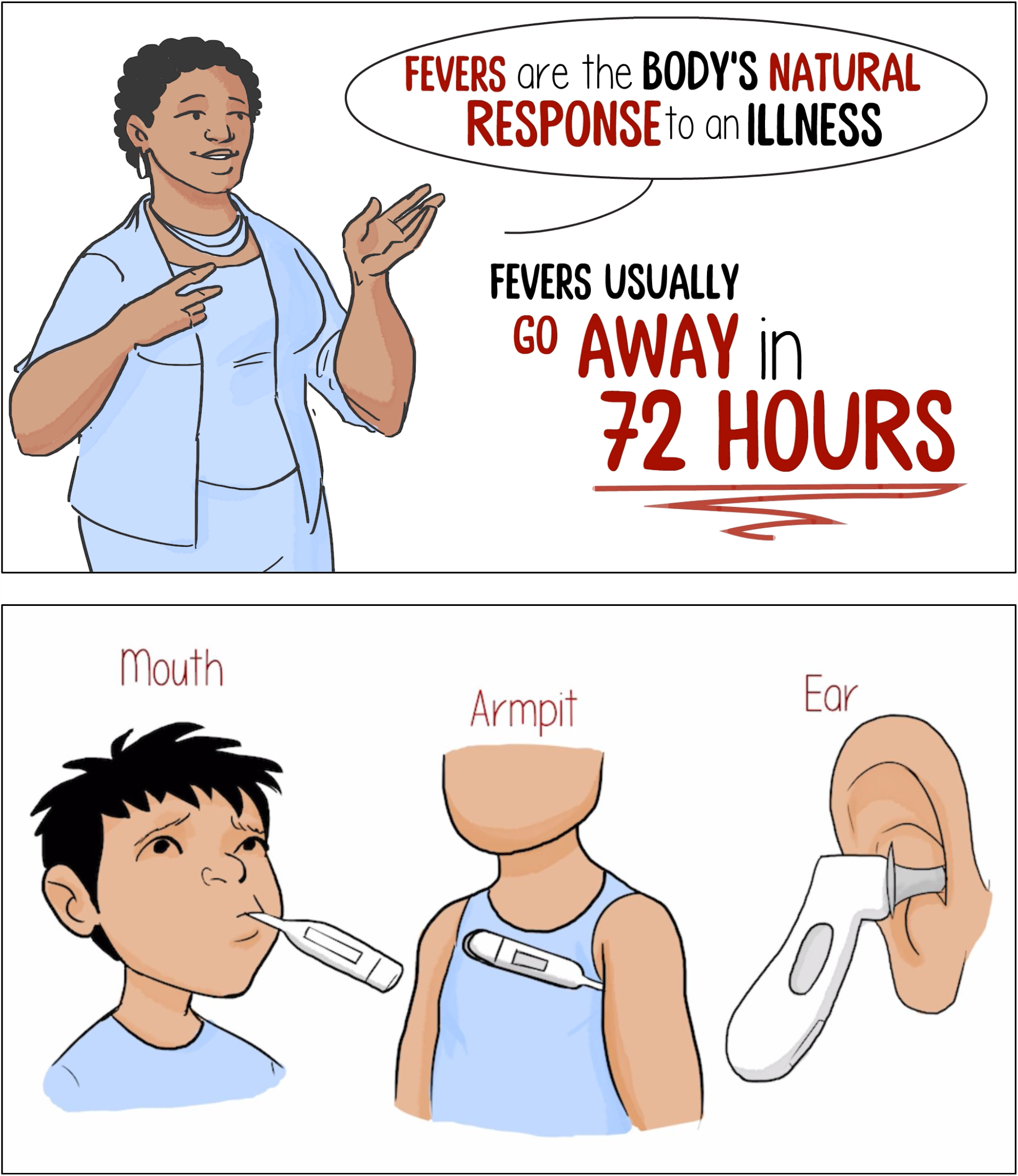

### Appendix C – Infographic

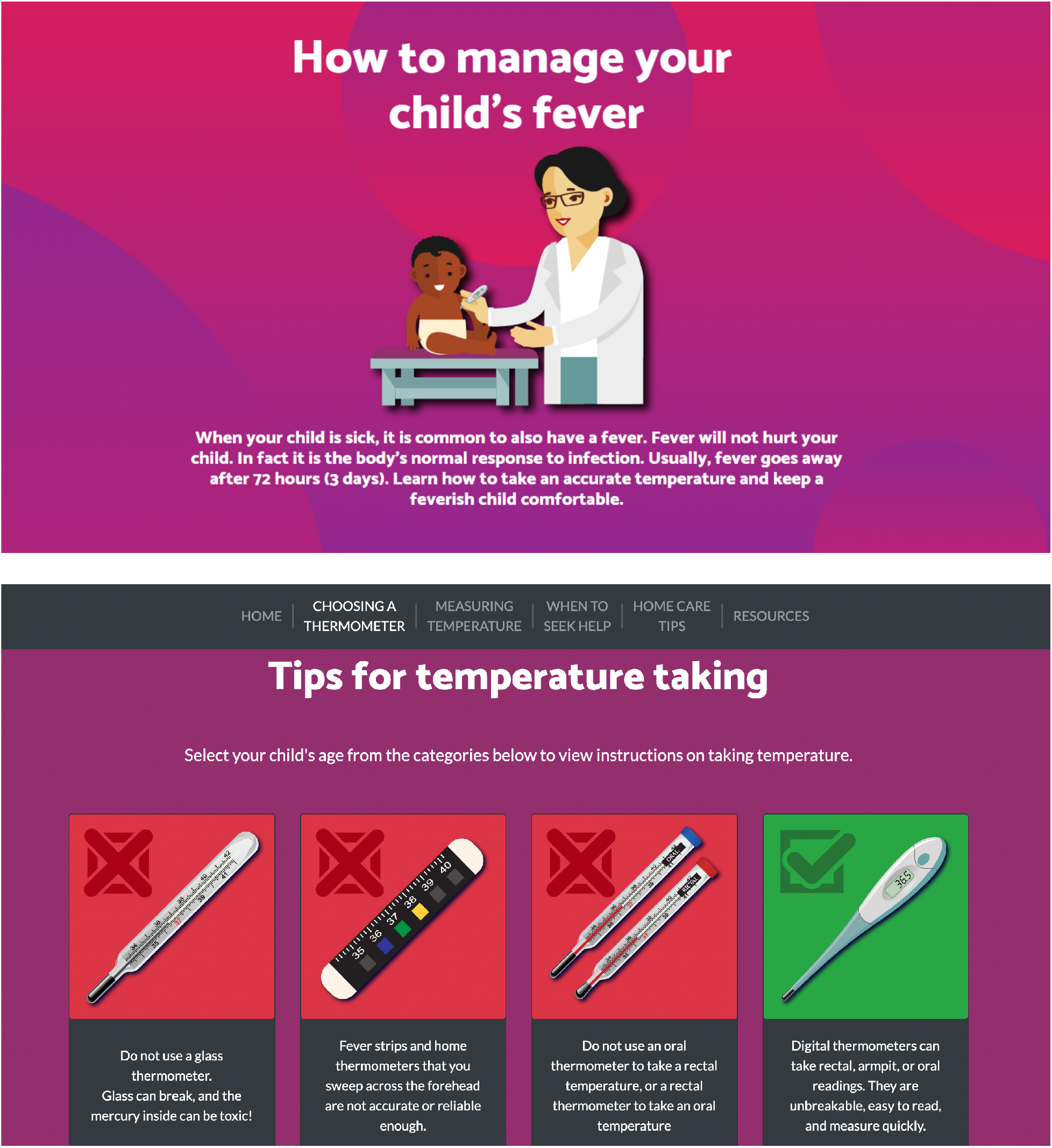

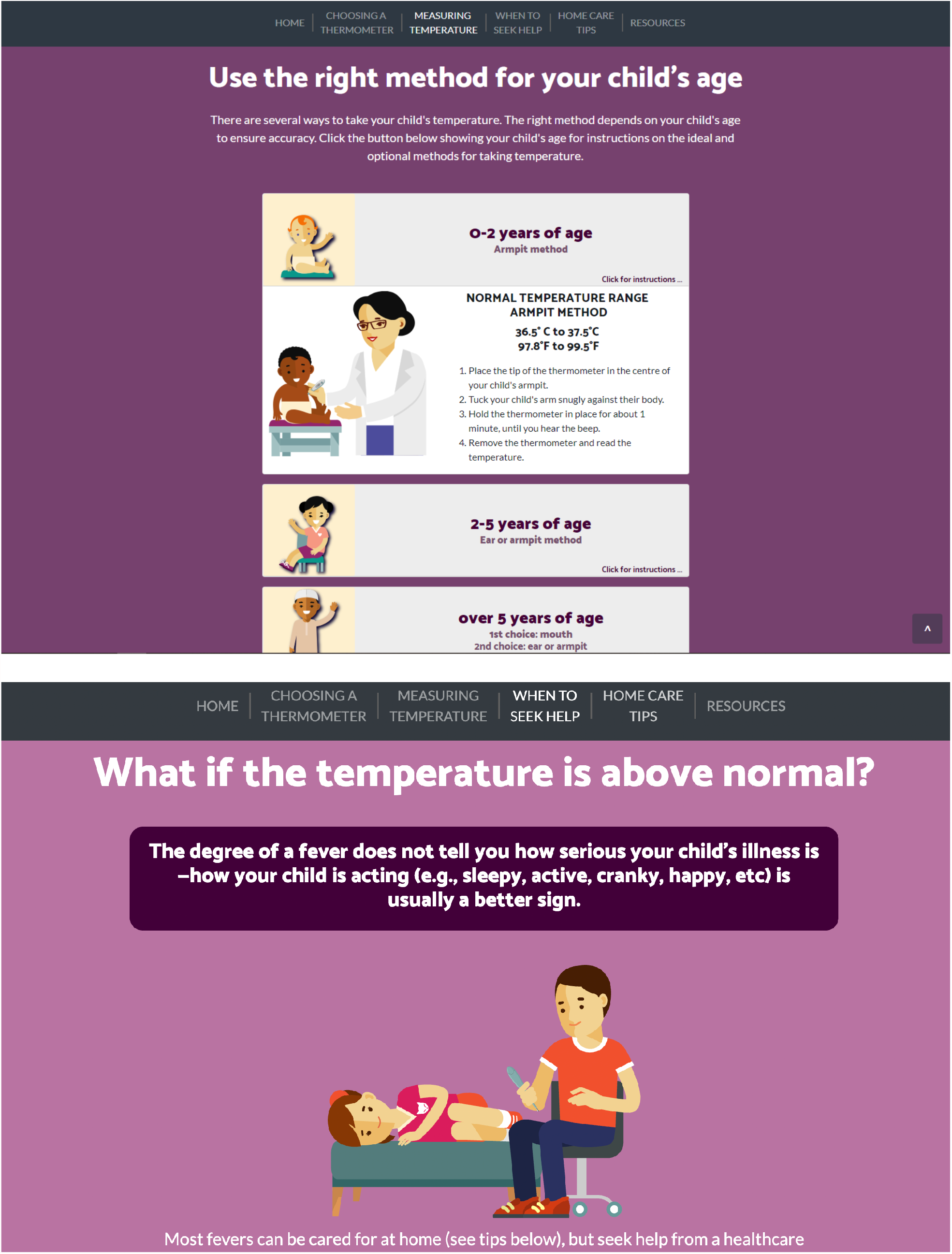

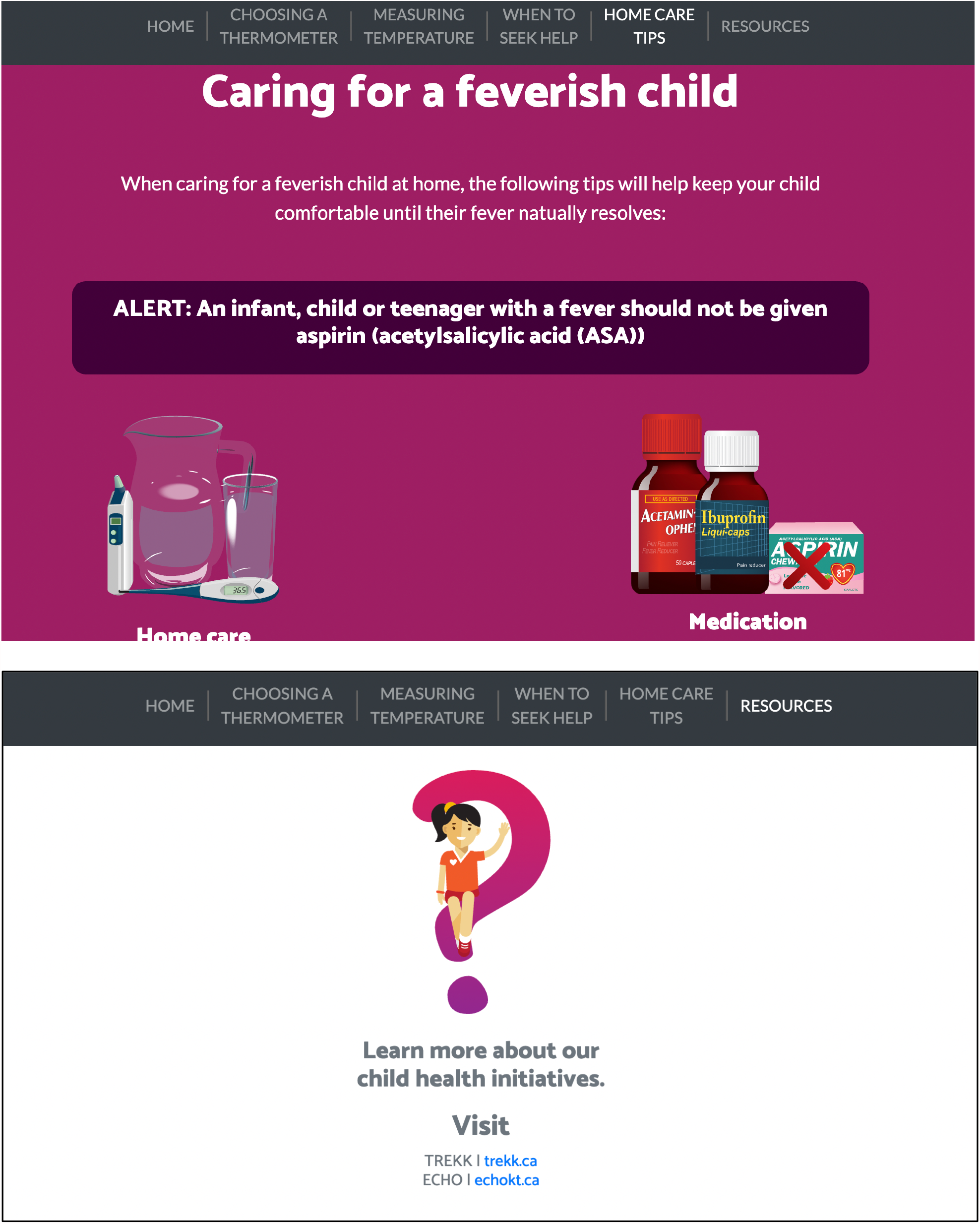

### Appendix D – Usability Survey

#### SECTION 1: Demographics

1) What is your gender?
  □ Male
  □ Female
3) What is your Age?
  □ Less than 20 years old
  □ 20-30 years
  □ 31-40 years
  □ 41-50 years
  □ 51 years and older
4) What is your Marital Status?
  □ Married
  □ Single
5) What is your gross annual household income?
  □ Less than $25,000
  □ $25,000-$49,999
  □ $50,000-$74,999
  □ $75,000-$99,999
  □ $100,000-$149,999
  □ $150,000 and over
6) What is your highest level of education?
  □ Some high school
  □ High school diploma
  □ Some post-secondary
  □ Post-secondary certificate/diploma
  □ Post-secondary degree
  □ Graduate degree
  □ Other
7) How many children do you have? _______
8) How old are your children? _______________

#### SECTION 2: Assessment of attributes of the arts-based, digital tools

***\*participant is randomized to view 1 of 2 digital tools then automatically directed to the survey***

1. It is useful. [5-point Likert Scale]
2. It provides information that is relevant to me as a parent. [5-point Likert Scale]
3. It is simple to use. [5-point Likert Scale]
4. I can use it without written instructions or additional help. [5-point Likert Scale]
5. Its length is appropriate. [5-point Likert Scale]
6. It is aesthetically pleasing (i.e., images, colours, etc.). [5-point Likert Scale]
7. It helps me to make decisions about my child’s health. [5-point Likert Scale]
8. I would use it in the future. [5-point Likert Scale]
9. I would recommend it to a friend. [5-point Likert Scale]
10. List the most negative aspects: [open text]
11. List the most positive aspects: [open text]

### Appendix E – Project Timeline

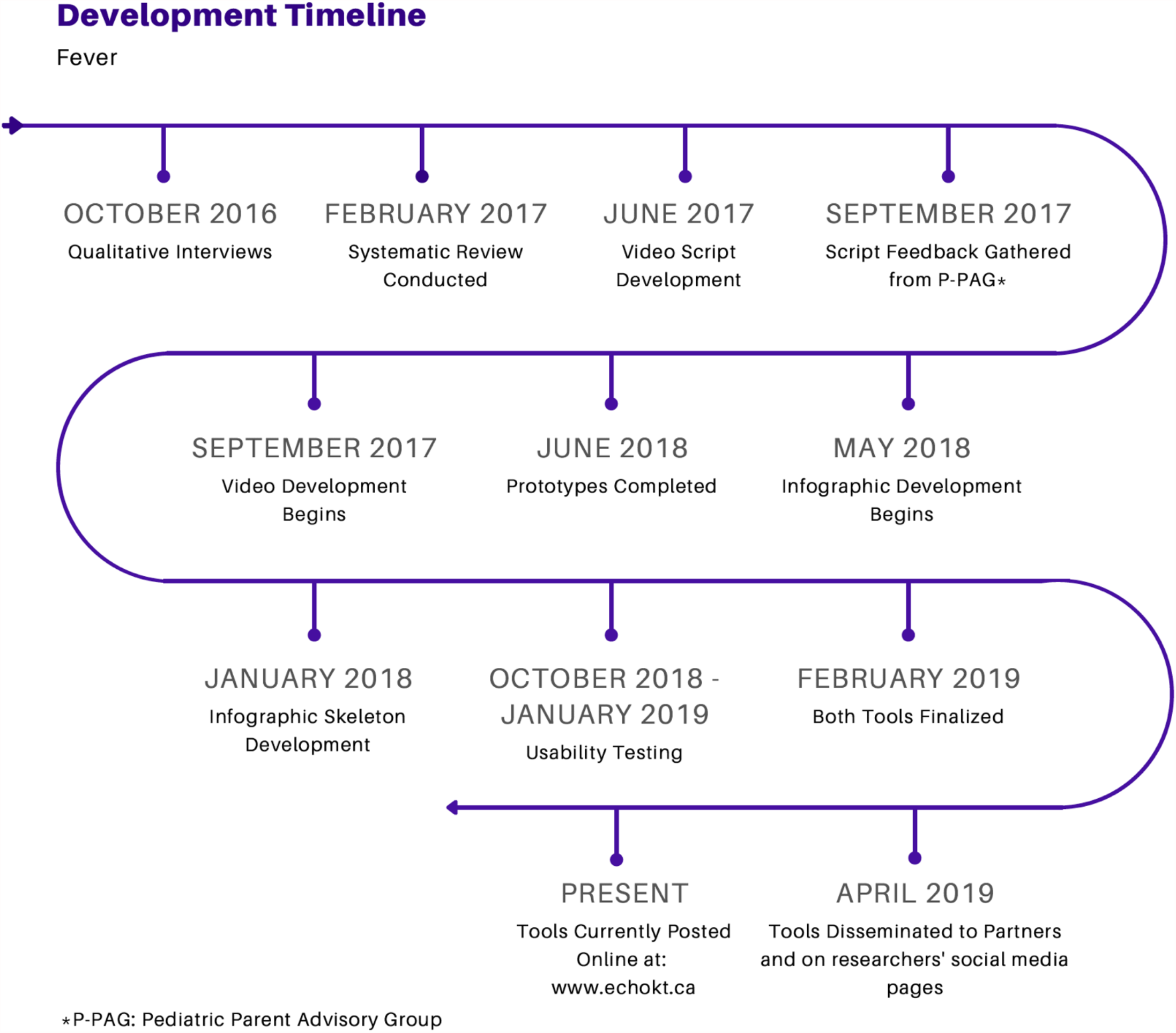

## Author Contributions

This study was conducted under the supervision of Drs. Shannon D. Scott (SDS) and Lisa Hartling (LH), PIs for **translation Evidence in Child Health to enhance Outcomes** (ECHO) Research and the **Alberta Research Centre for Health Evidence**, respectively. Both PIs designed the research study and obtained research funding through Translating Emergency Knowledge for Kids (TREKK) Networks of Centres of Excellence of Canada (NCE), Stollery Children’s Hospital Foundation, and the Women and Children’s Health Research Institute.

SDS led and supervised all aspects of tool development and evaluation.

Mary Klute conducted qualitative interviews with parents.

Alison Thompson, Mary Klute and Anne Le analyzed the qualitative interviews.

Chentel Cunningham and Alyson Campbell conducted usability testing.

AL and CC analyzed usability testing data.

CC developed the first draft of this technical report. All authors provided substantial feedback.

This work was funded by:

### Networks of Centres of Excellence

- Klassen, T., Hartling, L., Jabbour, M., Johnson, D., & Scott, S.D. (2015). Translating emergency knowledge for kids (TREKK). Networks of Centres of Excellence of Canada Knowledge Mobilization Renewal ($1,200,000). January 2016 – December 2019.

### Women’s and Children Health Research Institute

- Scott, S.D & Hartling L. (2016). Translating Emergency Knowledge for Kids renewal. Women and Children’s Health Research Institute (matched dollars, $150,000). 2016/04-2019/04.

### This report should be cited as

Scott, S.D., Cunningham, C., Le, A., Hartling, L. (2021). Development and usability testing of two arts-based knowledge translation tools for parents about pediatric fever. Internal Technical Report. ECHO Research, University of Alberta.

*Available at: http://www.echokt.ca/research/technical-reports/*

